# Racial/ethnic differences in pre-pregnancy conditions and adverse maternal outcomes in the nuMoM2b cohort: a population-based cohort study

**DOI:** 10.1101/2022.11.02.22281812

**Authors:** Meghan E. Meredith, Lauren N. Steimle, Kaitlyn K. Stanhope, Marissa H. Platner, Sheree L. Boulet

## Abstract

**Objective:** To: (1) determine how pre-existing conditions contribute to racial disparities in adverse maternal outcomes; and (2) incorporate these conditions into models to improve risk prediction for racial minority subgroups.

**Design:** Secondary data analysis of a population-based cohort study.

**Setting:** Academic healthcare institutions in the United States.

**Population:** A cohort of 8,729 women included in the “Nulliparous Pregnancy Outcomes Study: Monitoring Mothers-to-be (nuMoM2b)”.

**Methods:** Poisson regression to estimate adjusted risk ratios, and mediation analysis to evaluate the contribution of multimorbidity to racial/ethnic disparities. Area under the receiver operating characteristic curve (AUC) to evaluate model performance.

**Main Outcome Measures:** Incidence of severe preeclampsia, postpartum readmission, and blood transfusion.

**Results:** In the nuMoM2b cohort (n=8729), accounting for pre-existing conditions attenuated the association between non-Hispanic Black race/ethnicity and risk of severe preeclampsia. Cardiovascular and kidney conditions were associated with risk for severe preeclampsia among all women (aRR, 1.77; CI, 1.61-1.96, and aRR, 1.27; CI, 1.03-1.56 respectively). The mediation analysis results were not statistically significant; however, cardiovascular conditions explained 36.6% of the association between severe preeclampsia and non-Hispanic Black race/ethnicity (p=0.07). The addition of pre-pregnancy conditions increased model performance for the prediction of blood transfusion and severe preeclampsia.

**Conclusions:** Pre-existing conditions may explain some of the association between severe preeclampsia and non-Hispanic Black race/ethnicity. Specific pre-pregnancy conditions were associated with adverse maternal outcomes and the incorporation of comorbidities improved the performance of most risk prediction models.

**Funding:** Research reported in this publication was supported in part by Imagine, Innovate and Impact (I^3^) from the Emory School of Medicine, Georgia Tech, and through the Georgia CTSA NIH award (UL1-TR002378) and by the National Science Foundation under grant number DGE-2039655 (Meredith); any opinions, findings, and conclusions or recommendations expressed in this material are those of the authors and do not necessarily reflect the views of the National Science Foundation.

## Introduction

The global maternal mortality rate has stalled after improving between 2000 and 2015.^1^ The United States’ maternal mortality rate, 23.8 per 100,000 live births, has worsened over the past 20 years,^2,3^ and there are staggering racial and ethnic disparities in maternal mortality.^4^ In the United States, compared with non-Hispanic white individuals, non-Hispanic Black individuals have 3 to 4 times the risk of dying during childbirth.^5^ An estimated 80% of pregnancy-related deaths are considered preventable,^6^ and individuals with adverse maternal outcomes have similar preventable factors including provider failure to identify high-risk status and inappropriate management.^7^ Two potential targets for addressing pregnancy-related mortality and morbidity are improvements in chronic condition prevention before and during pregnancy and better prediction models for adverse outcomes that may disproportionally affect racial and ethnic minority individuals.

Chronic conditions are associated with a higher risk of adverse maternal outcomes. Deliveries among racial and ethnic minorities, compared with non-Hispanic white individuals, experience significantly higher rates of maternal morbidity and also have a higher prevalence of chronic conditions.^8^ Pregnant individuals with multiple co-occurring conditions, or *multimorbidity*, have higher rates of severe maternal morbidity and postpartum readmission.^9^ While multimorbidity affects a substantial and likely growing proportion of the global adult population,^10^ obstetric research and practice remain largely focused on the impact of single conditions on maternal outcomes,^11^ apart from a few studies using administrative data, ^12,13^ and those creating comorbidity-based risk screening tools.^14–16^ When multimorbidity is accounted for, it is typically represented as a binary indication of each condition or an indicator of the number of conditions, but these representations do not consider the types of co-occurring conditions (e.g. hematological and cardiovascular), which may be more relevant for clinical management.

In this study, we investigate the impact of pre-pregnancy conditions and their combined effects on adverse maternal outcomes and whether these effects mediate the relationship between racial disparities and adverse maternal outcomes in a cohort of nulliparous individuals in the United States. A better understanding of which conditions and combinations thereof drive increased maternal risk could inform the development of new clinical care standards for multimorbid pregnant people and potentially improve the ability to risk-stratify pregnant people early in pregnancy.

## Methods

### Study Population

We conducted a secondary analysis of data from the nuMoM2b prospective cohort study of nulliparous women.^17,18^ The study recruited women from hospitals affiliated with eight clinical centers and collected data on each participant over the course of four study visits via in-clinic interviews, self-administered questionnaires, clinical measurements, and chart abstractions. The cohort included 9,289 women who consented to the release of their data.

### Data Preparation

For this secondary data analysis, we chose to exclude women who delivered at gestational age < 22 weeks or > 43 weeks and those with terminations and unknown outcomes. We also excluded women with missing race/ethnicity and pre-pregnancy conditions data. To address missing data among confounders, we created 5 imputed datasets using multivariate imputation by chained equations (MICE).19 Please see Appendix A for more details about the exclusion protocol and Appendix B.1 for more details about the data structure and preparation.

### Main Outcome Measures

The primary outcomes of this study were severe preeclampsia, blood transfusion, and postpartum readmission (up to 14 days). These outcomes were selected because they are strongly associated with maternal mortality and morbidity, ^20,21^ and they have high quality measurements in the nuMoM2b dataset. Appendix B.2 provides details about the definitions and collection of the primary outcomes in the nuMoM2b study.

We fit prediction models to each of these outcomes individually to preserve the interpretability of model results and compared results across each outcome.

### Pre-Pregnancy Conditions

Pre-pregnancy conditions were collected by the nuMoM2b study team at each of the first three visits through an in-person interview with the participant, who indicated the presence of each condition at any time throughout their life. The nuMoM2b study team reconciled this data with chart abstraction to finalize the indication of pre-pregnancy conditions.

The nuMoM2b study team collected 41 pre-pregnancy conditions, and we categorized them into 12 condition types according to the framework of Tang et al.^22^ (Appendix Table S3). We grouped the pre-pregnancy conditions based on similarities in treatments, clinical manifestation, or organization in the health care system. For example, we grouped cervical dysplasia, fibroids, and PCOS under the condition type “Gynecological” because they are conditions that affect the function of female reproductive organs and are treated by the same gynecologist specialty branch. The condition types included: Autoimmune, Cardiovascular, Endocrine, Gastrointestinal, Gynecological, Hematologic, Kidney, Lung, Mental, and Neurological. This classification was developed and validated by three of our authors (KS, SB, MP) with extensive medical and specifically obstetric knowledge.

Our analysis considered pairs of co-occurring pre-pregnancy condition types. For example, consider a patient with the following three pre-pregnancy conditions: anemia (hematologic), hypertension (cardiovascular), and migraine headaches (neurological). This patient was indicated to have the following co-occurring condition types: hematologic & cardiovascular, cardiovascular & hematologic, and hematologic & neurological.

### Confounders

We adjusted for confounding factors including maternal age, insurance, body mass index (BMI), and sociodemographic information including income and education. Each of these factors were collected during Visit 1, which occurred between gestational age 6 and 14 weeks.

### Statistical Analysis

We used Poisson regression models with robust standard errors to analyze the associations between pre-pregnancy condition types, self-reported race/ethnicity, and adverse maternal outcomes with the results reported as crude and adjusted risk ratios (RRs) and 95% confidence intervals (CIs). We used a class-weighting method which gives a higher priority to correctly classifying the subgroup of women who experienced an adverse maternal outcome (see Appendix B.3).

Potential mediators were selected for each adverse maternal outcome in a stepwise process to avoid multicollinearity. Each adverse maternal outcome was regressed by race/ethnicity, confounders, and all potential mediators (all pre-pregnancy condition types and all their combined effects) (hereafter referred to as the “Outcome Model”). Only condition types and combined effects reaching statistical significance (p-value ≤ 0.05) in this model were included in further analysis. Additionally, only combined effects with at least 1% prevalence in the dataset were considered. Next, we used a model-building approach starting with a standard individual-level Poisson model (Model 1, crude), followed by a Poisson model that adjusted for confounders (Model 2, adjusted). We additionally controlled for significant pre-pregnancy condition types (Model 3), and finally, we additionally controlled for significant combined effects between co-occurring pre-pregnancy condition types (Model 4).

Next, we sought to determine if pre-pregnancy condition types and their combined effects contributed to the racial disparities in adverse maternal outcomes using mediation analysis. To analyze mediation, the potential mediator condition types and their combined effects were individually regressed by race/ethnicity and adjusted for confounders (“Mediator Model”). The Outcome and Mediator models were combined to compute the mediation proportion, which estimates the proportion of the risk factor’s impact (race/ethnicity) on the outcome that is attributable to the mediator (pre-pregnancy condition type or combined effect).^23^ This analysis was conducted for each adverse maternal outcome and results were averaged across the five imputed datasets.

### Performance Analysis

Finally, we analyzed the performance of our prediction models (Model 1, Model 2, Model 3, and Model 4) to understand which pre-pregnancy patient characteristics accurately classified adverse maternal outcomes. The performance of each model was evaluated for each racial/ethnic subgroup using the average area under the ROC curve (AUC) across 10-fold cross-validation and across the five imputed datasets.

We used R, version 4.2.1, for all analysis, *caret* in R to train and test our prediction models and *mediate* in R to conduct mediation analysis. We use a p-value threshold of 0.05 for statistical significance.

## Results

After exclusions, the final study population included 8729 women (Table 1). Of those, 61.0% (n=5322) identified as non-Hispanic white, 13.1% (n=1145) non-Hispanic Black, 17.0% (n=1487) Hispanic, 3.9% (n = 342) Asian, 4.0% (n=348) multiracial, and 1.0% (n=85) “Other”. The non-Hispanic Black subgroup had the youngest average age (23.3± 5.3) and the largest average BMI (29.1 ± 8.2). In total, the study population included 157 cases (1.8%) of blood transfusion, 154 cases (1.8%) of postpartum readmission, and 318 cases (3.6%) of severe preeclampsia. The incidence of these adverse maternal outcomes varied by race and ethnicity (Table Appendix S2). The incidence of blood transfusion and severe preeclampsia was highest in non-Hispanic Black women and the incidence of all adverse maternal outcomes was lowest in Asian women.

**Table 1:** Descriptive Statistics of the nuMoM2b Study Population by race/ethnicity status (before imputing missing data). BMI, body mass index; Std, standard deviation; HS, high school

|  | Asian | Hispanic | Multiracial | Non-Hispanic Black | Non-Hispanic White | Other |
| --- | --- | --- | --- | --- | --- | --- |
|  | N (%) | N (%) | N (%) | N (%) | N (%) | N (%) |
| **Total** | 342 (3.9%) | 1487 (17.0%) | 348 (4.0%) | 1145 (13.1%) | 5322 (61.0%) | 85 (1.0%) |
| **Education** |  |  |  |  |  |  |
| Degree work beyond college | 158 (46.2%) | 148 (10.0%) | 56 (16.1%) | 73 (6.4%) | 1546 (29.1%) | 23 (27.1%) |
| Completed college | 110 (32.2%) | 249 (16.8%) | 68 (19.5%) | 117 (10.2%) | 1835 (34.5%) | 26 (30.6%) |
| Some college | 31 (9.1%) | 443 (29.8%) | 87 (25.0%) | 336 (29.3%) | 795 (14.9%) | 12 (14.1%) |
| Assoc/Tech degree | 29 (8.5%) | 165 (11.1%) | 29 (8.3%) | 114 (10.0%) | 543 (10.2%) | 7 (8.2%) |
| HS grad or GED | 9 (2.6%) | 271 (18.2%) | 47 (13.5%) | 291 (25.4%) | 391 (7.4%) | 10 (11.8%) |
| Less than HS grad | 4 (1.2%) | 206 (13.9%) | 61 (17.5%) | 214 (18.7%) | 212 (4.0%) | 6 (7.1%) |
| Missing | 1 (0.3%) | 5 (0.3%) | 0 (0%) | 0 (0%) | 0 (0%) | 1 (1.2%) |
| **Gravidity** |  |  |  |  |  |  |
| 1 | 262 (76.6%) | 1014 (68.2%) | 250 (71.8%) | 798 (69.7%) | 4116 (77.3%) | 63 (74.1%) |
| 2 | 61 (17.8%) | 334 (22.5%) | 77 (22.1%) | 226 (19.7%) | 937 (17.6%) | 16 (18.8%) |
| 3 + | 19 (5.6%) | 139 (9.4%) | 21 (6.0%) | 121 (10.6%) | 269 (5.1%) | 6 (7.1%) |
| **Insurance** |  |  |  |  |  |  |
| Government | 34 (9.9%) | 825 (55.5%) | 147 (42.2%) | 692 (60.4%) | 764 (14.4%) | 25 (29.4%) |
| Commercial | 289 (84.5%) | 593 (39.9%) | 178 (51.2%) | 383 (33.5%) | 4406 (82.8%) | 54 (63.5%) |
| Self-pay | 86 (25.2%) | 144 (9.7%) | 47 (13.5%) | 71 (6.2%) | 1130 (21.2%) | 15 (17.7%) |
| Other | 8 (2.3%) | 31 (2.1%) | 11 (3.2%) | 34 (3.0%) | 87 (1.6%) | 2 (2.4%) |
| **Type of Labor** |  |  |  |  |  |  |
| Spontaneous, not augmented | 69 (20.2%) | 270 (18.2%) | 67 (19.3%) | 168 (14.7%) | 953 (17.9%) | 15 (17.7%) |
| Spontaneous, augmented | 164 (48.0%) | 650 (43.7%) | 130 (37.4%) | 462 (40.4%) | 2181 (41.0%) | 32 (37.7%) |
| Induced | 81 (23.7%) | 475 (31.9%) | 131 (37.6%) | 463 (40.4%) | 1808 (34.0%) | 33 (38.8%) |
| Cesarean w/o labor or induction | 28 (8.2%) | 91 (6.1%) | 20 (5.8%) | 51 (4.5%) | 379 (7.1%) | 5 (5.9%) |
| **Mode of Delivery** |  |  |  |  |  |  |
| Vaginal | 234 (68.4%) | 1046 (70.3%) | 264 (75.9%) | 788 (68.8%) | 3923 (73.7%) | 55 (64.7%) |
| Cesarean |  |  |  |  |  |  |
| Planned or scheduled | 19 (5.6%) | 60 (4.0%) | 11 (3.2%) | 32 (2.8%) | 282 (5.3%) | 3 (3.5%) |
| Unscheduled, non-urgent | 49 (14.3%) | 188 (12.6%) | 44 (12.6%) | 181 (15.8%) | 702 (13.2%) | 21 (24.7%) |
| Unscheduled, urgent | 40 (11.7%) | 193 (13.0%) | 29 (8.3%) | 144 (12.6%) | 411 (7.7%) | 6 (7.1%) |
|  | Mean ± Std | Mean ± Std | Mean ± Std | Mean ± Std | Mean ± Std | Mean ± Std |
| **Age** | 31.0 ± 4.7 | 24.8 ± 5.52 | 25.0 ± 6.2 | 23.3 ± 5.3 | 28.2 ± 5.2 | 27.9 ± 5.1 |
| **BMI** | 23.3 ± 4.0 | 26.7 ± 6.0 | 27.1 ± 6.9 | 29.1 ± 8.2 | 25.8 ± 5.8 | 27.2 ± 6.3 |
| **Percent Federal Poverty** | 590.9 ± 315.8 | 293.3 ± 263.2 | 337.4 ± 295.1 | 208.0 ± 245.0 | 488.3 ± 299.8 | 394.3 ± 287.1 |
| **Gestational Age at Delivery (weeks)** | 38.9 ± 1.9 | 38.8 ± 2.0 | 38.6 ± 2.1 | 38.4 ± 2.5 | 38.9 ± 2.0 | 38.4 ± 2.6 |

The most common pre-pregnancy conditions were mental health conditions (14.4%, n=1261), hematologic conditions (13.8%, n=1206), neurological conditions (13.0%, n=1136), and lung conditions (12.5%, n=1091) (Table Appendix S4). Non-Hispanic Black women experienced the highest rates of lung and hematologic conditions, Hispanic women experienced the highest rates of kidney conditions, and multiracial women experienced the highest rates of cardiovascular conditions. Non-Hispanic white women experienced the highest rates of neurological, gastrointestinal, and mental health conditions.

Of all women, 22.8% (n=1989) had co-occurring pre-pregnancy conditions, i.e., 2 or more conditions. Non-Hispanic white women had the highest rates of co-occurring pre-pregnancy conditions (24.9%, n=1327), followed by multiracial women (22.7%, n=79), and Non-Hispanic Black women (20.0%, n=229). The most common co-occurring pre-pregnancy conditions were Mental & Neurological (3.7%, n=322), Lung & Neurological (2.8%, n=248), and Hematologic & Lung (2.5%, n=216) (Table Appendix S5). Among non-Hispanic white women, the most common co-occurring pre-pregnancy conditions were Mental & Neurological (4.7%, n=248), Lung & Neurological (2.9%, n=153), and Hematologic & Lung (2.1%, n=111). Among non-Hispanic Black women, the most common co-occurring pre-pregnancy conditions were Hematologic & Lung (4.4%, n=50) and Cardiovascular & Hematologic (3.4%, n=39).

Compared with non-Hispanic white women, the unadjusted risk of blood transfusion and severe preeclampsia was higher in non-Hispanic Black women. Statistical adjustment for age, sociodemographic information, insurance, and BMI eliminated the elevated risk of blood transfusion in non-Hispanic Black and Hispanic women and eliminated the elevated risk of severe preeclampsia for multiracial women. Statistical adjustment attenuated but did not eliminate the elevated risk of severe preeclampsia in non-Hispanic Black women. The adjusted relative risk (aRR) of severe preeclampsia was higher in non-Hispanic Black (aRR, 1.22; CI, 1.06-1.41) compared to non-Hispanic white women (Table 4).

After adjusting for confounders, we controlled for pre-pregnancy condition types (Model 3) and their combined effects (Model 4). Controlling for pre-pregnancy condition types for the outcome of severe preeclampsia attenuated the risk of severe preeclampsia in non-Hispanic Black women (aRR 1.15, CI, 0.99-1.34) (Table 6).

Pre-pregnancy condition types were associated with adverse maternal outcomes (see Tables 2-4). Autoimmune conditions significantly increased the risk for blood transfusion (aRR, 1.36; CI, 1.04-1.78) and postpartum readmission (aRR, 1.55; CI, 1.22-1.97). Hematologic conditions were associated with a significantly increased risk for blood transfusion (aRR, 1.42; CI, 1.23-1.64). Cardiovascular and kidney conditions were associated with a significantly increased risk for severe preeclampsia (aRR, 1.77; CI, 1.61-1.96, and aRR, 1.27; CI, 1.03-1.56 respectively). Accounting for combined effects did not significantly impact effect estimates.

**Table 2:** Multilevel Poisson regression with outcome blood transfusion. Bolding indicates statistical significance at 95% confidence. RR, risk ratio; CI, confidence interval; AUC, area under the receiver operating characteristic curve

|  | Model 1, crude | Model 2, adjusted | Model 3, condition types<br>RR (95% CI) | Model 4, combined effects |
| --- | --- | --- | --- | --- |
| Race |  |  |  |  |
| Non-Hispanic White (reference) | 1.00 | 1.00 | 1.00 | 1.00 |
| Non-Hispanic Black | **1.29 ( 1.06 - 1.57 )** | 1.20 ( 0.97 - 1.50 ) | 1.19 ( 0.95 - 1.50 ) | 1.21 ( 0.96 - 1.52 ) |
| Hispanic | **1.28 ( 1.07 - 1.54 )** | 1.17 ( 0.95 - 1.45 ) | 1.19 ( 0.96 - 1.48 ) | 1.19 ( 0.96 - 1.48 ) |
| Multiracial | 0.98 ( 0.59 - 1.62 ) | 0.94 ( 0.57 - 1.52 ) | 0.86 ( 0.54 - 1.36 ) | 0.84 ( 0.53 - 1.32 ) |
| Asian | 0.86 ( 0.47 - 1.59 ) | 0.85 ( 0.46 - 1.57 ) | 0.88 ( 0.48 - 1.63 ) | 0.88 ( 0.48 - 1.63 ) |
| Other | 1.25 ( 0.67 - 2.32 ) | 1.21 ( 0.66 - 2.22 ) | 1.17 ( 0.66 - 2.06 ) | 1.18 ( 0.67 - 2.09 ) |
| Confounders |  |  |  |  |
| Pre-Pregnancy Conditions |  |  |  |  |
| Cardiovascular |  |  | 1.11 ( 0.88 - 1.39 ) | 1.17 ( 0.92 - 1.48 ) |
| Endocrine |  |  | 1.25 ( 1.00 - 1.56 ) | **1.28 ( 1.02 - 1.60 )** |
| Kidney |  |  | 1.28 ( 0.93 - 1.75 ) | 1.27 ( 0.93 - 1.75 ) |
| Hematologic |  |  | **1.42 ( 1.23 - 1.64 )** | **1.42 ( 1.23 - 1.64 )** |
| Autoimmune |  |  | **1.36 ( 1.04 - 1.78 )** | **1.36 ( 1.04 - 1.79 )** |
| Gynecological |  |  | 0.69 ( 0.48 - 1.00 ) | 0.73 ( 0.50 - 1.06 ) |
| Combined effects |  |  |  |  |
| Cardiovascular: Endocrine |  |  |  | 0.83 ( 0.38 - 1.83 ) |
| Cardiovascular: Gynecological |  |  |  | 0.67 ( 0.22 - 2.02 ) |
| Overall AUC | 0.56 | 0.55 | 0.59 | 0.59 |

**Table 3:**
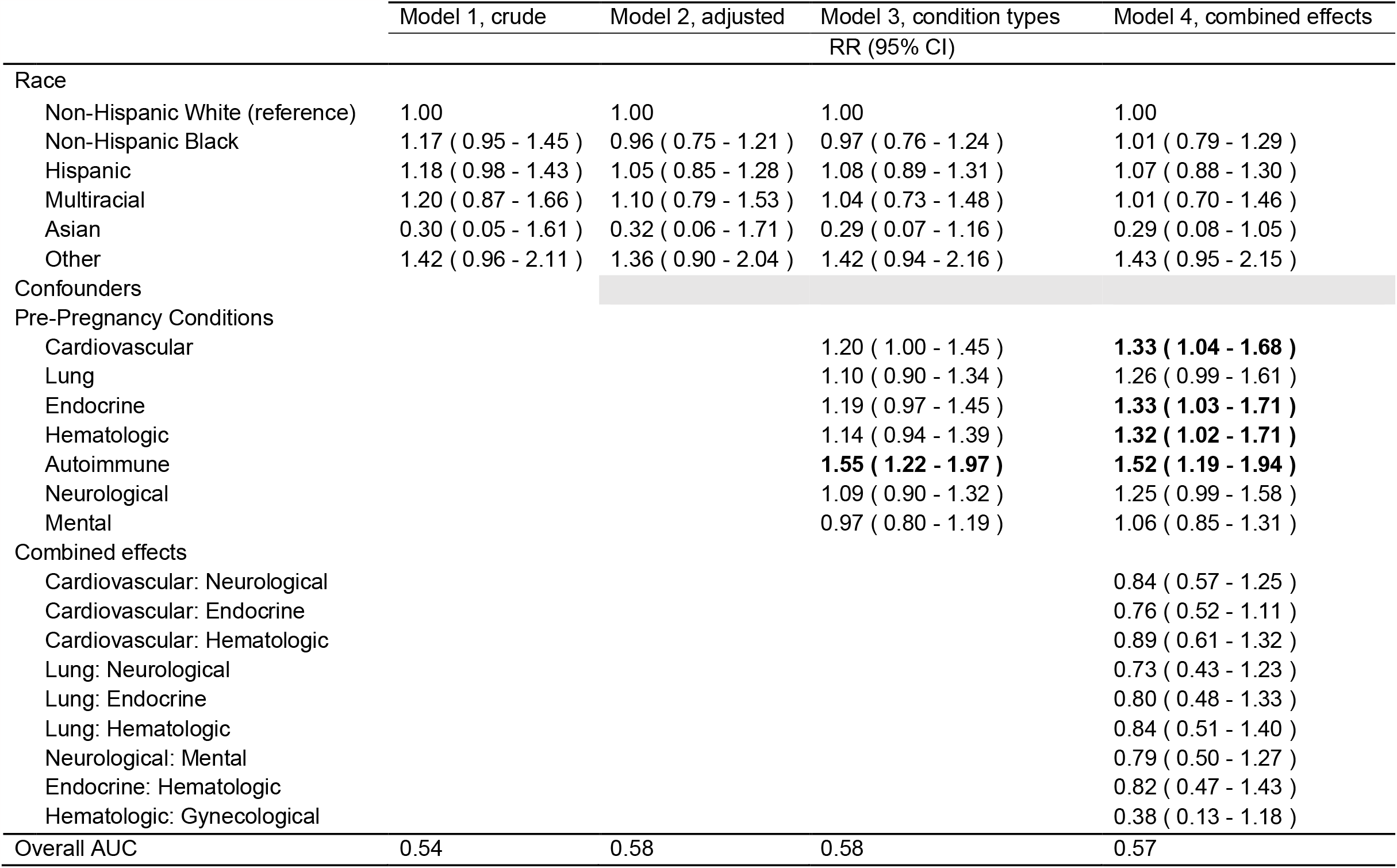
Multilevel Poisson regression with outcome postpartum readmission. Bolding indicates statistical significance at 95% confidence. RR, risk ratio; CI, confidence interval; AUC, area under the receiver operating characteristic curve

|  | Model 1, crude | Model 2, adjusted | Model 3, condition types | Model 4, combined effects |
| --- | --- | --- | --- | --- |
|  | RR (95% CI) |  |  |  |
| Race |  |  |  |  |
| Non-Hispanic White (reference) | 1.00 | 1.00 | 1.00 | 1.00 |
| Non-Hispanic Black | 1.17 ( 0.95 - 1.45 ) | 0.96 ( 0.75 - 1.21 ) | 0.97 ( 0.76 - 1.24 ) | 1.01 ( 0.79 - 1.29 ) |
| Hispanic | 1.18 ( 0.98 - 1.43 ) | 1.05 ( 0.85 - 1.28 ) | 1.08 ( 0.89 - 1.31 ) | 1.07 ( 0.88 - 1.30 ) |
| Multiracial | 1.20 ( 0.87 - 1.66 ) | 1.10 ( 0.79 - 1.53 ) | 1.04 ( 0.73 - 1.48 ) | 1.01 ( 0.70 - 1.46 ) |
| Asian | 0.30 ( 0.05 - 1.61 ) | 0.32 ( 0.06 - 1.71 ) | 0.29 ( 0.07 - 1.16 ) | 0.29 ( 0.08 - 1.05 ) |
| Other | 1.42 ( 0.96 - 2.11 ) | 1.36 ( 0.90 - 2.04 ) | 1.42 ( 0.94 - 2.16 ) | 1.43 ( 0.95 - 2.15 ) |
| Confounders |  |  |  |  |
| Pre-Pregnancy Conditions |  |  |  |  |
| Cardiovascular |  |  | 1.20 ( 1.00 - 1.45 ) | **1.33 ( 1.04 - 1.68 )** |
| Lung |  |  | 1.10 ( 0.90 - 1.34 ) | 1.26 ( 0.99 - 1.61 ) |
| Endocrine |  |  | 1.19 ( 0.97 - 1.45 ) | **1.33 ( 1.03 - 1.71 )** |
| Hematologic |  |  | 1.14 ( 0.94 - 1.39 ) | **1.32 ( 1.02 - 1.71 )** |
| Autoimmune |  |  | **1.55 ( 1.22 - 1.97 )** | **1.52 ( 1.19 - 1.94 )** |
| Neurological |  |  | 1.09 ( 0.90 - 1.32 ) | 1.25 ( 0.99 - 1.58 ) |
| Mental |  |  | 0.97 ( 0.80 - 1.19 ) | 1.06 ( 0.85 - 1.31 ) |
| Combined effects |  |  |  |  |
| Cardiovascular: Neurological |  |  |  | 0.84 ( 0.57 - 1.25 ) |
| Cardiovascular: Endocrine |  |  |  | 0.76 ( 0.52 - 1.11 ) |
| Cardiovascular: Hematologic |  |  |  | 0.89 ( 0.61 - 1.32 ) |
| Lung: Neurological |  |  |  | 0.73 ( 0.43 - 1.23 ) |
| Lung: Endocrine |  |  |  | 0.80 ( 0.48 - 1.33 ) |
| Lung: Hematologic |  |  |  | 0.84 ( 0.51 - 1.40 ) |
| Neurological: Mental |  |  |  | 0.79 ( 0.50 - 1.27 ) |
| Endocrine: Hematologic |  |  |  | 0.82 ( 0.47 - 1.43 ) |
| Hematologic: Gynecological |  |  |  | 0.38 ( 0.13 - 1.18 ) |
| Overall AUC | 0.54 | 0.58 | 0.58 | 0.57 |

**Table 4:** Multilevel Poisson regression with outcome severe preeclampsia, bolding indicates statistical significance at 95% confidence. RR, risk ratio; CI, confidence interval; AUC, area under the receiver operating characteristic curve

|  | Model 1, crude | Model 2, adjusted | Model 3, condition types | Model 4, combined effects |
| --- | --- | --- | --- | --- |
|  | RR (95% CI) |  |  |  |
| (Intercept) | **0.45 ( 0.41 - 0.49 )** | **0.23 ( 0.16 - 0.33 )** | **0.23 ( 0.16 - 0.34 )** | **0.23 ( 0.16 - 0.34 )** |
| Race |  |  |  |  |
| Non-Hispanic White (reference) | 1.00 | 1.00 | 1.00 | 1.00 |
| Non-Hispanic Black | **1.45 ( 1.29 - 1.63 )** | **1.22 ( 1.06 - 1.41 )** | 1.15 ( 0.99 - 1.34 ) | 1.16 ( 0.99 - 1.35 ) |
| Hispanic | 1.09 ( 0.92 - 1.28 ) | 1.04 ( 0.87 - 1.23 ) | 0.97 ( 0.82 - 1.14 ) | 0.97 ( 0.82 - 1.15 ) |
| Multiracial | **1.28 ( 1.02 - 1.60 )** | 1.09 ( 0.88 - 1.35 ) | 1.06 ( 0.85 - 1.32 ) | 1.06 ( 0.85 - 1.31 ) |
| Asian | 0.93 ( 0.62 - 1.37 ) | 0.98 ( 0.67 - 1.43 ) | 0.96 ( 0.67 - 1.38 ) | 0.97 ( 0.67 - 1.38 ) |
| Other | 1.26 ( 0.81 - 1.96 ) | 1.20 ( 0.78 - 1.86 ) | 1.21 ( 0.76 - 1.91 ) | 1.21 ( 0.77 - 1.91 ) |
| Confounders |  |  |  |  |
| Pre-Pregnancy Conditions |  |  |  |  |
| Cardiovascular |  |  | **1.77 ( 1.61 - 1.96 )** | **1.71 ( 1.54 - 1.89 )** |
| Kidney |  |  | **1.27 ( 1.03 - 1.56 )** | **1.25 ( 1.01 - 1.53 )** |
| Hematologic |  |  | 0.90 ( 0.77 - 1.04 ) | 0.80 ( 0.59 - 1.08 ) |
| Gastrointestinal |  |  | 0.71 ( 0.48 - 1.05 ) | 0.70 ( 0.47 - 1.05 ) |
| Gynecological |  |  | 0.84 ( 0.70 - 1.02 ) | 0.85 ( 0.71 - 1.02 ) |
| Combined effects |  |  |  |  |
| Cardiovascular: Hematologic |  |  |  | 1.26 ( 0.91 - 1.75 ) |
| Overall AUC | 0.56 | 0.61 | 0.65 | 0.65 |

Mediation analysis was conducted on all statistically significant condition types and combined effects for each adverse maternal outcome. There were no significant results at p-value < 0.05; however, cardiovascular conditions accounted for 36.6% of the association between non-Hispanic Black race/ethnicity and severe preeclampsia at p-value 0.07.

Finally, we compared the performance metrics, specifically the area under the receiver operating characteristics curve (AUC), of models with different feature sets to determine the value of different maternal characteristics in predicting the risk of experiencing each of the adverse maternal outcomes (Table Appendix S6). We found gains in overall AUC with the addition of condition types for the prediction of blood transfusion and severe preeclampsia (AUC, 0.55 to 0.59, 0.61 to 0.65, respectively). We computed AUC for each racial and ethnic subgroup to understand how the models performed across the subgroups. AUC varies significantly by race for each adverse maternal outcome. Model 3, which includes race, confounders, and condition types as features, has an AUC range of 0.50-0.67 for blood transfusion, 0.45-0.63 for postpartum readmission, and 0.49-0.70 for severe preeclampsia. The largest improvement in AUC from adding condition was in predicting blood transfusion in Multiracial women, from 0.49 to 0.67.

## Discussion

### Main Findings

Our findings suggest accounting for pre-pregnancy condition types explained some of the association between non-Hispanic Black race/ethnicity and severe preeclampsia. We found that including pre-pregnancy condition types improved diagnostic ability (AUC) in predictive models for blood transfusion and severe preeclampsia, both for the overall cohort and for most individual racial and ethnic groups. These results highlight the potential for risk prediction using pre-pregnancy conditions in a diverse, but low-risk population.

Consistent with previous studies,^5,8,24–26^ we found that identification as a minority race was associated with a higher risk of adverse maternal outcomes among a cohort of nulliparous individuals. Adding to some studies that have examined the impact of specific co-occurring condition combined effects,^27,28^ our study focused on exploring all potential pre-pregnancy condition type combined effects to understand their association with adverse maternal outcomes (specifically severe preeclampsia, postpartum readmission, and blood transfusion) and quantified the value of including combined effects in predictive models. No pre-pregnancy condition combined effects were significantly associated with adverse maternal outcomes.

We also quantified the association between individual pre-pregnancy condition type and adverse maternal outcomes. Each adverse maternal outcome was associated with different pre-pregnancy condition types, except the autoimmune condition type, which was associated with increased risk for both blood transfusion and postpartum readmission. It is well known that pregnancy is associated with changes in the maternal immune system, and specifically the postpartum period is associated with autoimmune disease flares.^29^

Our work adds to previous studies on the use of prediction models to predict various adverse outcomes, which have ranged from using symptoms and signs,^14,16^ laboratory tests and biomarkers,^30,31^ and demographics and medical history.^32,33^ With the inclusion of pre-pregnancy condition types, our model yields an AUC of 0.65 for predicting severe preeclampsia, which is comparable to the AUC of previously published obstetric comorbidity indices predicting SMM, which included severe preeclampsia (Bateman, et al.^15^: AUC, 0.65 and Easter, et al.^14^ : AUC, 0.70 as reported by Leonard et al.^34^). The prediction models for blood transfusion and severe preeclampsia improved with the addition of pre-pregnancy condition types; however, the model for postpartum readmission did not improve.

### Interpretation

Understanding the potential reasons for adverse maternal outcomes is an important pathway to understanding and reducing racial and ethnic disparities and high rates of maternal morbidity and mortality.

Our findings suggest that the presence of pre-pregnancy conditions and confounders may explain some of the observed association between non-Hispanic Black race/ethnicity and severe preeclampsia. In a prior study, Black race was associated with increased odds of pregnancy-induced hypertension after adjustment for preexisting conditions and demographic factors.^35^ However, our study adjusts for a different set of preexisting conditions. The prevention and management of these pre-existing conditions, specifically cardiovascular conditions, before, during, and between pregnancies could be an important consideration to decrease racial and ethnic disparities in adverse maternal outcomes. Although the nuMoM2b cohort study collected data on a wide range of pre-pregnancy conditions, we could not explain a large proportion of the disparities in maternal morbidity in this cohort. Future work may benefit from the inclusion of additional data such as the severity and management of morbidity and multimorbidity during the preconception, interconception, and postpartum periods to determine whether differential management reduces the observed disparities.

Our work suggests that information about pre-pregnancy conditions can be useful in improving the ability to risk-stratify individuals. The use of predictive modeling may be useful to further explore the complex relationships of co-occurring conditions. However, more research is necessary to inform best clinical practice for the use of predictive models with a focus on mitigating unintended consequences and preventing the exacerbation of disparities.^36^

### Strengths and Limitations

Our study has several strengths. Using a large and comprehensive dataset, we evaluated the association between race and ethnicity, pre-pregnancy conditions, and adverse maternal outcomes in a cohort of nulliparous women in the United States. This dataset contains thoroughly collected data that goes beyond a typical electronic health record including health history and conditions, demographics, and survey questionnaires. Our condition type groupings allowed for larger sample sizes of conditions and more accurate estimates of risk ratios. In addition, the use of feature selection algorithms allowed for the exploration of combined effects to improve model performance for predicting adverse maternal outcomes.

Our study has several important limitations. First, the analysis was limited to the data collected in the nuMoM2b dataset, which only includes nulliparous, predominantly non-Hispanic white women who received regular prenatal care at academic medical centers and began care before 13 weeks and 6 days gestation; thus, our findings may not be generalizable to different birthing populations. Second, the nuMoM2b dataset also does not indicate the severity or long-term treatment status of pre-pregnancy conditions. Third, our dataset only collected data up to 14 days postpartum, although many postpartum readmissions occur after this timespan.^37^ Finally, adverse pregnancy outcomes are rare and occur infrequently in the nuMoM2b dataset, particularly as we stratified by racial subgroups and by multimorbidity. A larger sample size may be able to identify more statistically significant associations between pre-pregnancy conditions and maternal outcomes that occur infrequently.

## Conclusion

In addition to describing associations between race and ethnicity, pre-pregnancy condition types and their combined effects, and adverse maternal outcomes, our findings indicate that pre-pregnancy conditions may partially explain the association between severe preeclampsia and non-Hispanic Black race/ethnicity. Additionally, data collected at an initial prenatal care visit has utility for predicting the risk of experiencing an adverse maternal outcome. Our study findings have important implications for the preconception care as well as antepartum care of individuals with pre-existing conditions as adequately assessing a patient’s risk is essential to providing risk-appropriate and equitable care to prevent adverse maternal outcomes.

## Supporting information

Supplemental maternal

## Data Availability

The data is not available to be shared per our data use agreement.

## Acknowledgments

None.

## Disclosure of Interests

The authors report no disclosures of interest.

## Contribution to Authorship

All authors were involved in the literature search and in the design of the study. MM and LNS obtained the data and MM performed the data analysis. MM drafted the manuscript with LNS. KS, MP, and SH revised the manuscript for important intellectual content, and all authors were involved in the interpretation of the results. All authors approved the final version to be published.

## Details of Ethics Approval

This study was approved by the Georgia Institute of Technology Internal Review Board which deemed it exempt from review.

---

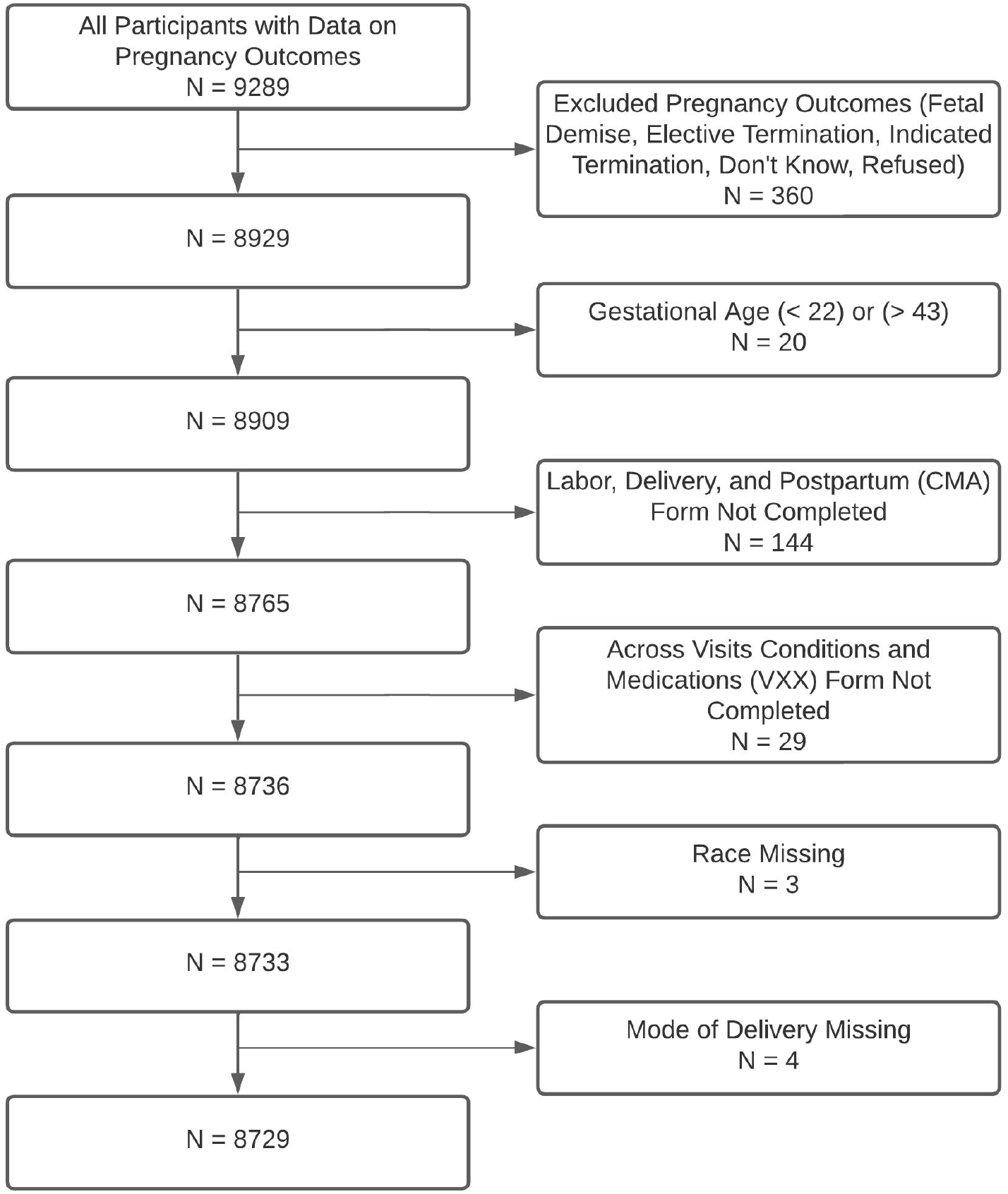

## Notes

### Competing Interest Statement

The authors have declared no competing interest.

### Funding Statement

Research reported in this publication was supported in part by Imagine, Innovate and Impact (I3) from the Emory School of Medicine, Georgia Tech, and through the Georgia CTSA NIH award (UL1-TR002378) and by the National Science Foundation under grant number DGE-2039655 (Meredith); any opinions, findings, and conclusions or recommendations expressed in this material are those of the authors and do not necessarily reflect the views of the National Science Foundation.

### Author Declarations

This study was approved by the Georgia Institute of Technology Internal Review Board who deemed it exempt from review

### Summary of Updates

Updated to clarify findings.

