## Supplemental maternal for "Racial/ethnic differences in pre-pregnancy conditions and adverse maternal outcomes in the nuMoM2b cohort: a population-based cohort study"

### Supporting Information

#### Appendix A. Exclusion Protocol

For this secondary data analysis, we chose to exclude women with pregnancy outcomes including fetal death before 20 weeks gestation, elective termination, indicated termination, unknown outcomes, and if the participant refused to release their pregnancy outcome. We also excluded women who delivered at gestational age < 22 weeks or > 43 weeks. We excluded women who had an incomplete labor, delivery, and postpartum form (form “CMA” in the nuMoM2b database) and those with an incomplete across visit medical conditions and medications form (form “VXX” in the nuMoM2b database).[18] Finally, we excluded women without self-reported race/ethnicity status and those without mode of delivery recorded (Figure S1).

Figure S1: Exclusion Criteria

### Appendix B: Methods

#### B.1 Data Preparation

The nuMoM2b study recruited individuals from hospitals affiliated with eight clinical centers and collected data on each participant over the course of four study visits. Visit 1 occurred between 6 and 14 weeks, Visit 2 occurred between 16 and 22 weeks, Visit 3 occurred between 22 and 30 weeks, and Visit 4 occurred at the time of delivery.

In the nuMoM2b study, women's racial and ethnic status was self-reported during Visit 1, and women were given the option to select more than one racial category. In our analysis, we used a single feature to represent race and ethnicity. We grouped maternal race and ethnicity as non-Hispanic white, non-Hispanic Black, Hispanic, Asian, multiracial, and other, where the "Other" category includes American Indian, Native Hawaiian, and any other race/ethnicity status.

No features were missing at more than 20%, so none were removed. We imputed the missing entries using multivariate imputation by chained equations (MICE),[38] an algorithm that preserves the distribution of each variable. Specifically, we used predictive mean matching (PMM)[38] because this method imputes only values that are observed and therefore preserves categorical variables.

#### B.2 Primary Outcomes

The outcome of "severe preeclampsia" in our study is indicated by one of the following diagnoses from the nuMoM2b dataset: severe preeclampsia, superimposed preeclampsia, or eclampsia. These diagnoses were identified through a detailed chart review performed by a site investigator or staff member certified for abstraction of complicated charts. If necessary, difficult cases were adjudicated by the principal investigators and final classification was reached by consensus. The outcome of "blood transfusion" is indicated by any amount of any blood product administered anytime during pregnancy or postpartum (up to 14 days) as recorded in a participant's medical records. The occurrence of postpartum readmission is indicated by any readmission to the hospital within 14 days of delivery. A detailed description of the nuMoM2b study definitions of hypertensive orders is included as a supplement to the paper by Facco et al. (2017).[39]

Table S1: Adverse maternal outcomes and their corresponding indicators in the nuMoM2b dataset

| **Adverse Maternal Outcome** | **nuMoM2b Variable** | **Description** |
|---|---|---|
| Severe Preeclampsia Superimposed Eclampsia | PEgHTN | Preeclampsia/Gestational hypertension (HTN) (worst) using nuMoM2b criteria (CMDA08a) |
| Blood Product Transfusion | CMAE11 | Any blood product transfusion anytime during pregnancy through 14 days postpartum |
| Postpartum Readmission | CMAJ01 | Participant readmitted to the hospital within 14 days of delivery |

Table S2: Rates of adverse maternal outcomes in the nuMoM2b population overall and by race/ethnicity

| | Asian (N=342) | Hispanic (N=1487) | Multiracial (N=348) | Non-Hispanic Black (N=1145) | Non-Hispanic White (N=5322) | Other (N=85) | Overall (N=8729) |
|---|---|---|---|---|---|---|---|
| **Blood Transfusion** | | | | | | | |
| Yes | 4 (1.2%) | 37 (2.5%) | 5 (1.4%) | 29 (2.5%) | 80 (1.5%) | 2 (2.4%) | 157 (1.8%) |
| No | 338 (98.8%) | 1449 (97.4%) | 343 (98.6%) | 1116 (97.5%) | 5242 (98.5%) | 83 (97.6%) | 8571 (98.2%) |
| Missing | 0 (0%) | 1 (0.1%) | 0 (0%) | 0 (0%) | 0 (0%) | 0 (0%) | 1 (0.0%) |
| **Postpartum Readmission** | | | | | | | |
| Yes | 1 (0.3%) | 33 (2.2%) | 8 (2.3%) | 25 (2.2%) | 84 (1.6%) | 3 (3.5%) | 154 (1.8%) |
| No | 341 (99.7%) | 1451 (97.6%) | 340 (97.7%) | 1119 (97.7%) | 5230 (98.3%) | 82 (96.5%) | 8563 (98.1%) |
| Missing | 0 (0%) | 3 (0.2%) | 0 (0%) | 1 (0.1%) | 8 (0.2%) | 0 (0%) | 12 (0.1%) |
| **Severe Preeclampsia** | | | | | | | |
| Yes | 9 (2.6%) | 52 (3.5%) | 17 (4.9%) | 76 (6.6%) | 160 (3.0%) | 4 (4.7%) | 318 (3.6%) |
| No | 332 (97.1%) | 1431 (96.2%) | 331 (95.1%) | 1069 (93.4%) | 5156 (96.9%) | 81 (95.3%) | 8400 (96.2%) |
| Missing | 1 (0.3%) | 4 (0.3%) | 0 (0%) | 0 (0%) | 6 (0.1%) | 0 (0%) | 11 (0.1%) |

#### B.3 Study variables from the nuMoM2b dataset

Table S3: Pre-pregnancy conditions and their corresponding indicators in the nuMoM2b dataset

| Condition Type | Pre-Pregnancy Condition | nuMoM2b Variable |
|---|---|---|
| Acute Infection | Urinary tract infection | VXXB01am_V1a |
| Acute Infection | Sexually transmitted diseases, Gonorrhea | VXXB01bd1_V1a |
| Acute Infection | Sexually transmitted diseases, Chlamydia | VXXB01bd2_V1a |
| Acute Infection | Sexually transmitted diseases, Herpes | VXXB01bd3_V1a |
| Acute Infection | Sexually transmitted diseases, Syphilis | VXXB01bd4_V1a |
| Acute Infection | Sexually transmitted diseases, HIV/AIDS | VXXB01bd5_V1a |
| Acute Infection | Sexually transmitted diseases, Hepatitis B | VXXB01bd6_V1a |
| Acute Infection | Sexually transmitted diseases, Hepatitis C | VXXB01bd7_V1a |
| Acute Infection | Sexually transmitted diseases, Trichomonas | VXXB01bd8_V1a |
| Acute Infection | Sexually transmitted diseases, Other | VXXB01bd9_V1a |
| Acute Infection | Yeast infection | VXXB01be_V1a |
| Acute Infection | Bacterial vaginosis | VXXB01bf_V1a |
| Autoimmune | Systemic lupus erythematosus (SLE) | VXXB01au_V1a |
| Autoimmune | Rheumatoid arthritis | VXXB01av_V1a |
| Autoimmune | Other collagen vascular or autoimmune disease | VXXB01aw_V1a |
| Cancer | Cancer (malignancy) | VXXB01ba_V1a |
| Cardiovascular | High blood pressure (hypertension) | VXXB01aa_V1a |
| Cardiovascular | Valvular heart disease | VXXB01ah_V1a |
| Cardiovascular | Other structural heart disease | VXXB01ai_V1a |
| Cardiovascular | Coronary artery disease / congestive heart failure | VXXB01aj_V1a |
| Cardiovascular | Cardiac arrhythmias | VXXB01ak_V1a |
| Endocrine | Diabetes (excluding gestational diabetes in a prior pregnancy) | VXXB01ae_V1a |
| Endocrine | Hyperthyroidism | VXXB01af_V1a |
| Endocrine | Hypothyroidism | VXXB01ag_V1a |
| Gastrointestinal | Ulcerative colitis / Crohn's disease | VXXB01ax_V1a |
| Gastrointestinal | Liver/gall bladder disease | VXXB01az_V1a |
| Gynecological | Cervical dysplasia | VXXB01bc1_V1a |
| Gynecological | Fibroids | VXXB01bc2_V1a |
| Gynecological | Polycystic ovary disease (PCOS) | VXXB01bc3_V1a |
| Hematologic | Sickle cell disease | VXXB01an_V1a |
| Hematologic | Sickle cell trait | VXXB01ao_V1a |

| | | |
|---|---|---|
| Hematologic | Thrombocytopenia | VXXB01ap_V1a |
| Hematologic | Anemia | VXXB01aq_V1a |
| Hematologic | History of blood clots (thrombosis or thromboembolism) or stroke | VXXB01ar_V1a |
| Hematologic | Congenital or inherited bleeding disorder | VXXB01as_V1a |
| Hematologic | Antiphospholipid syndrome (APA) or other acquired thrombophilia | VXXB01at_V1a |
| Kidney | Kidney disease | VXXB01al_V1a |
| Lung | Asthma | VXXB01ab_V1a |
| Mental | Mental health condition | VXXB01bb_V1a |
| Neurological | Seizure disorder | VXXB01ac_V1a |
| Neurological | Migraine headaches | VXXB01ad_V1a |

In our analysis, we excluded the condition types: Acute Infection and Cancer. Acute infection is recorded as occurring anytime during a participant’s life, which lacks relevance in our analysis because the nuMoM2b study did not collect the timing of the acute infection and because lifetime acute infections are common. Cancer was excluded due to its low prevalence.

Table S4: Prevalence of pre-pregnancy conditions by race in nuMoM2b dataset

Number of condition types is the sum of different condition types, for example, if a woman only has 2 cardiovascular conditions this is counted as 1 condition type

| | Asian | Hispanic | Multiracial | Non-Hispanic Black | Non-Hispanic White | Other | Overall |
|---|---|---|---|---|---|---|---|
| **Total** | 342 (3.9%) | 1487 (17.0%) | 348 (4.0%) | 1145 (13.1%) | 5322 (61.0%) | 85 (1.0%) | 8729 (100.0%) |
| **Cardiovascular** | 20 (5.8%) | 102 (6.9%) | 43 (12.4%) | 120 (10.5%) | 459 (8.6%) | 5 (5.9%) | 749 (8.6%) |
| **Lung** | 31 (9.1%) | 174 (11.7%) | 53 (15.2%) | 181 (15.8%) | 640 (12.0%) | 12 (14.1%) | 1091 (12.5%) |
| **Neurological** | 31 (9.1%) | 165 (11.1%) | 37 (10.6%) | 116 (10.1%) | 778 (14.6%) | 9 (10.6%) | 1136 (13.0%) |
| **Endocrine** | 40 (11.7%) | 109 (7.3%) | 15 (4.3%) | 65 (5.7%) | 494 (9.3%) | 10 (11.8%) | 733 (8.4%) |
| **Kidney** | 2 (0.6%) | 41 (2.8%) | 2 (0.6%) | 12 (1.0%) | 97 (1.8%) | 2 (2.4%) | 156 (1.8%) |
| **Hematologic** | 42 (12.3%) | 223 (15.0%) | 63 (18.1%) | 221 (19.3%) | 647 (12.2%) | 10 (11.8%) | 1206 (13.8%) |
| **Autoimmune** | 6 (1.8%) | 19 (1.3%) | 5 (1.4%) | 18 (1.6%) | 121 (2.3%) | 2 (2.4%) | 171 (2.0%) |
| **Gastrointestinal** | 5 (1.5%) | 28 (1.9%) | 7 (2.0%) | 14 (1.2%) | 116 (2.2%) | 1 (1.2%) | 171 (2.0%) |
| **Mental** | 16 (4.7%) | 144 (9.7%) | 45 (12.9%) | 95 (8.3%) | 953 (17.9%) | 8 (9.4%) | 1261 (14.4%) |
| **Gynecological** | 48 (14.0%) | 126 (8.5%) | 30 (8.6%) | 82 (7.2%) | 591 (11.1%) | 8 (9.4%) | 885 (10.1%) |
| **Number of Condition Types** | | | | | | | |
| 0 | 181 (52.9%) | 769 (51.7%) | 161 (46.3%) | 597 (52.1%) | 2443 (45.9%) | 44 (51.8%) | 4195 (48.1%) |
| 1 | 105 (30.7%) | 435 (29.3%) | 108 (31.0%) | 319 (27.9%) | 1552 (29.2%) | 26 (30.6%) | 2545 (29.2%) |
| 2+ | 56 (16.4%) | 283 (19.0%) | 79 (22.7%) | 229 (20.0%) | 1327 (24.9%) | 15 (17.6%) | 1989 (22.8%) |

Table S5: Prevalence of pre-pregnancy condition combined effects included in our analysis by race in nuMoM2b dataset

| | Asian | Hispanic | Multiracial | Non-Hispanic Black | Non-Hispanic White | Other | Overall |
|---|---|---|---|---|---|---|---|
| **Total** | 342 (3.9%) | 1487 (17.0%) | 348 (4.0%) | 1145 (13.1%) | 5322 (61.0%) | 85 (1.0%) | 8729 (100.0%) |
| **Cardiovascular & Endocrine** | 2 (0.6%) | 12 (0.8%) | 2 (0.6%) | 27 (2.4%) | 64 (1.2%) | 2 (2.4%) | 109 (1.2%) |
| **Cardiovascular & Gynecological** | 3 (0.9%) | 10 (0.7%) | 5 (1.4%) | 20 (1.7%) | 70 (1.3%) | 1 (1.2%) | 109 (1.2%) |
| **Cardiovascular & Hematologic** | 2 (0.6%) | 22 (1.5%) | 12 (3.4%) | 39 (3.4%) | 84 (1.6%) | 2 (2.4%) | 161 (1.8%) |
| **Cardiovascular & Neurological** | 2 (0.6%) | 20 (1.3%) | 9 (2.6%) | 21 (1.8%) | 99 (1.9%) | 2 (2.4%) | 153 (1.8%) |
| **Endocrine & Hematologic** | 6 (1.8%) | 23 (1.5%) | 1 (0.3%) | 22 (1.9%) | 69 (1.3%) | 1 (1.2%) | 122 (1.4%) |
| **Endocrine & Lung** | 3 (0.9%) | 18 (1.2%) | 2 (0.6%) | 18 (1.6%) | 83 (1.6%) | 1 (1.2%) | 125 (1.4%) |
| **Gynecological & Hematologic** | 8 (2.3%) | 26 (1.7%) | 6 (1.7%) | 33 (2.9%) | 89 (1.7%) | 1 (1.2%) | 163 (1.9%) |
| **Hematologic & Lung** | 4 (1.2%) | 40 (2.7%) | 9 (2.6%) | 50 (4.4%) | 111 (2.1%) | 2 (2.4%) | 216 (2.5%) |
| **Lung & Neurological** | 8 (2.3%) | 37 (2.5%) | 11 (3.2%) | 35 (3.1%) | 153 (2.9%) | 4 (4.7%) | 248 (2.8%) |
| **Mental & Neurological** | 5 (1.5%) | 33 (2.2%) | 8 (2.3%) | 28 (2.4%) | 248 (4.7%) | 0 (0%) | 322 (3.7%) |

##### B.3 Model development details

The balancing class weight heuristic is inspired by King et al (2001) [40] and identical to scikit learn's "balanced" class weight function. The weights are given by the formula:

$$w_i = \frac{number\ of\ total\ samples}{(number\ of\ classes)\ \cdot\ (number\ of\ i^{th}\ outcome)}$$

Where,

$$i = \begin{cases} 1 & if\ adverse\ outcome\ occurred \\ 0 & otherwise \end{cases}$$

$number\ of\ total\ samples\ =\ 8729$

$number\ of\ classes\ =\ 2$

##### B.4 Mediation Analysis and Correlation

Firstly, each adverse maternal outcome was regressed individually by race/ethnicity, confounders, and potential mediators (all pre-pregnancy condition types and all their combined effects) (hereafter referred to as the "Outcome Model"), and only combined effects and their components reaching statistical significance (p-value ≤ 0.05) in this model were selected for further analysis. To analyze mediation, the selected potential mediator condition types and their combined effects were individually regressed by race/ethnicity and adjusted for confounders ("Mediator Model"). The Outcome and Mediator models were combined to compute average causal mediation effects (ACME) and total effect (TE) for each participant which was then averaged. Quasi-Bayesian estimation with 1,000 iterations was used to estimate the 95% CI and p-values of the natural indirect effect (NIE) and TE.[23] Mediation proportion, which estimates the proportion of the risk factor's impact on the outcome that is attributable to the mediator, was calculated as ACME / TE.[23] This analysis was conducted for each adverse maternal outcome and results were averaged across the five imputed datasets.

We assessed whether features were highly correlated and, a priori, planned to remove any that were highly correlated with another measure that was more relevant to our analysis to preserve the interpretability of the risk ratios. High correlation was indicated by a Pearson’s coefficient absolute value of 0.70 or greater. No features, other than confounding factors, were highly correlated.

#### Appendix C: Results

*Table S6: AUC by race for each model and adverse maternal outcome.*

*AUC, area under the receiver operating characteristic curve*

| | Model 0<br>Intercept | Model 1<br>Race | Model 2<br>(1) & Confounders | Model 3<br>(2) & Condition Types | Model 4<br>(3) & Condition Type Combined Effects |
|---|---|---|---|---|---|
| Blood Transfusion, AUC | | | | | |
| Non-Hispanic White | 0.49 | 0.50 | 0.51 | 0.58 | 0.58 |
| Hispanic | 0.49 | 0.50 | 0.50 | 0.50 | 0.51 |
| Non-Hispanic Black | 0.49 | 0.50 | 0.54 | 0.60 | 0.60 |
| Multiracial | 0.54 | 0.50 | 0.49 | 0.67 | 0.71 |
| Asian | 0.52 | 0.50 | 0.44 | 0.51 | 0.54 |
| Other | 0.58 | 0.90 | 0.63 | 0.52 | 0.52 |
| *Overall* | *0.49* | *0.56* | *0.55* | *0.59* | *0.59* |
| Postpartum Readmission, AUC | | | | | |
| Non-Hispanic White | 0.48 | 0.50 | 0.55 | 0.53 | 0.55 |
| Hispanic | 0.55 | 0.50 | 0.60 | 0.63 | 0.60 |
| Non-Hispanic Black | 0.52 | 0.50 | 0.59 | 0.59 | 0.57 |
| Multiracial | 0.57 | 0.55 | 0.53 | 0.59 | 0.54 |
| Asian | 0.76 | 0.50 | 0.50 | 0.50 | 0.50 |
| Other | 0.52 | 0.56 | 0.57 | 0.45 | 0.55 |
| *Overall* | *0.49* | *0.54* | *0.58* | *0.58* | *0.57* |
| Severe Preeclampsia, AUC | | | | | |
| Non-Hispanic White | 0.50 | 0.50 | 0.59 | 0.62 | 0.63 |
| Hispanic | 0.49 | 0.47 | 0.58 | 0.69 | 0.68 |
| Non-Hispanic Black | 0.51 | 0.50 | 0.52 | 0.58 | 0.58 |
| Multiracial | 0.47 | 0.50 | 0.71 | 0.70 | 0.72 |
| Asian | 0.50 | 0.50 | 0.65 | 0.69 | 0.69 |
| Other | 0.45 | 0.67 | 0.51 | 0.49 | 0.38 |
| *Overall* | *0.50* | *0.56* | *0.61* | *0.65* | *0.65* |
